# Prediction of Episode of Hemodynamic Instability Using an Electrocardiogram Based Analytic: A Retrospective Cohort Study

**DOI:** 10.1101/2023.06.08.23291138

**Authors:** Bryce Benson, Ashwin Belle, Sooin Lee, Benjamin S. Bassin, Richard P. Medlin, Michael W. Sjoding, Kevin R. Ward

## Abstract

**Background:** Predicting the onset of hemodynamic instability before it occurs remains a sought-after goal in acute and critical care medicine. Technologies that allow for this may assist clinicians in preventing episodes of hemodynamic instability (EHI). We tested a novel noninvasive technology, the Analytic for Hemodynamic Instability-Predictive Indicator (AHI-PI), which analyzes a single lead of electrocardiogram (ECG) and extracts heart rate variability and morphologic waveform features to predict an EHI prior to its occurrence.

**Methods:** Retrospective cohort study at a quaternary care academic health system using data from hospitalized adult patients between August 2019 and April 2020 undergoing continuous ECG monitoring with intermittent noninvasive blood pressure (NIBP) or with continuous intra-arterial pressure (IAP) monitoring.

**Results:** AHI-PI’s low and high-risk indications were compared with the presence of EHI in the future as indicated by vital signs (heart rate > 100 beats/min with a systolic blood pressure < 90 mmHg or a mean arterial blood pressure of < 70 mmHg). 4,633 patients were analyzed (3,961 undergoing NIBP monitoring, 672 with continuous IAP monitoring). 692 patients had an EHI (380 undergoing NIBP, 312 undergoing IAP). For IAP patients, the sensitivity and specificity of AHI-PI to predict EHI was 89.7% and 78.3% with a positive and negative predictive value of 33.7% and 98.4% respectively. For NIBP patients, AHI-PI had a sensitivity and specificity of 86.3% and 80.5% with a positive and negative predictive value of 11.7% and 99.5% respectively. Both groups performed with an AUC of 0.87. AHI-PI predicted EHI in both groups with a median lead time of 1.1 hours (average lead time of 3.7 hours for IAP group, 2.9 hours for NIBP group).

**Conclusions:** AHI-PI predicted EHIs with high sensitivity and specificity and within clinically significant time windows that may allow for intervention. Performance was similar in patients undergoing NIBP and IAP monitoring.

## Background

Sudden, unrecognized, and delayed identification of hemodynamic deterioration of patients remains a significant challenge across all levels of in-hospital care including the intensive care unit (ICU). Failure to recognize the need for re-evaluation and escalation of care can result in unplanned ICU admissions, added length of stay, and even death.^1–3^ This has prompted the development and implementation of a number of electronic medical record (EMR) deterioration indices or early warning systems (EWS) as well as rapid response teams that are designed to identify high risk patients in need of re-evaluation and to optimally intervene prior to deterioration. ^4–6^

While studies have demonstrated that shifts in vital signs can occur prior to adverse events and that close deliberate monitoring of vital signs may improve early detection and prompt clinical action capable of averting events such as cardiac arrest, high resolution monitoring, recording, and interpretation of vital signs is difficult even in the ICU setting.^7–9^ While several EMR technologies have demonstrated promise, the infrequency of EMR input (lab values, vital signs, etc.) and the subsequent validation of that input potentially reduces the effectiveness of such approaches to identify patients early.^10^

Loss of heart rate variability (HRV) has been demonstrated to reflect changes in the autonomic nervous system in the setting of many states of critical illness and injury including hemorrhage, sepsis, cardiogenic shock, respiratory failure, and others, with these changes occurring prior to overt decompensation.^11–17^ However, several challenges ranging from signal acquisition and sampling rates to real-time signal processing and feature extraction have limited the approach. A single lead ECG analytic was recently developed to overcome these challenges by leveraging advanced signal processing to extract HRV and ECG morphologic features associated with hemodynamic instability.^11, 18, 19^ The Analytic for Hemodynamic Instability-Predictive Indicator (AHI-PI) is designed to predict hemodynamic instability before it occurs. The analytic is an FDA cleared software as a medical device (SaMD). In this analysis, we report AHI-PI’s ability to predict a future occurrence of hemodynamic instability prior to it being identifiable by vital signs.

## Methods

This was a retrospective single-center observational cohort study conducted at a quaternary care academic health system. The study was approved by the University of Institutional Review Board using deidentified data (HUM00092309). The Institutional Review Board waived the need for informed consent since all data analyzed was retrospective and deidentified.

The study dataset included 4,633 consecutively hospitalized adult (≥18 years) patient encounters with continuous ECG monitoring between August 2019 and April 2020 across multiple levels of care including the emergency department, telemetry and step-down units, and ICUs. The University has a unique data acquisition and storage system that collects, stores, and maintains a high-resolution physiologic signal database of patients including real-time physiologic signals and waveforms such as ECG, arterial blood pressure, pulse oximetry and others.

AHI-PI utilizes streaming data from a single existing lead (II) of ECG to provide information regarding the patient’s predicted future risk for clinical deterioration based on the known physiologic relationship of HRV, the autonomic nervous system, and the cardiovascular system. AHI automates the extraction and analysis of ECG patterns that reflect the compensatory burden of the autonomic nervous system including signal quality assessment and processing of extracted patterns through a pretrained classification model that embeds nonlinear HRV complexity and ECG morphologic features into a signal output.^11, 18, 19^ AHI-PI builds on this output and updates every two minutes, producing one of three types of outputs: red, yellow, or green, indicating high, moderate, or low risk respectively of a future episode of hemodynamic instability.

In this analysis, an episode of hemodynamic instability (EHI) is defined as the presence of hypotension (systolic blood pressure <90 mmHg or mean arterial pressure <70 mmHg) combined with tachycardia (heart rate ≥ 100 bpm). This combination of blood pressure and heart rate to define EHI as it relates to inpatient adverse outcomes including mortality is supported by several widely used critical care and EWS systems including the National Early Warning Score (NEWS), the electronic Cardiac Arrest Triage (eCART), the Modified Early Warning Score (MEWS), and others.^4, 20–22^ Blood pressure was either taken by intermittent noninvasive blood pressure (NIBP) monitoring and recorded in the EMR after nurse validation or with continuous intraarterial blood pressure (IAP) monitoring. For patients undergoing IAP monitoring an EHI is defined more conservatively as 10 continuous minutes or more with the above heart rate and blood pressure parameters. For patients undergoing continuous IAP monitoring, blood pressure values and heart rate were collected at 0.5 Hz. However, patients monitored with NIBP had heart rate and blood pressure measures recorded approximately once per hour across all patients. Only ECG data was used as input for the AHI-PI algorithm, while the heart rate and blood pressure values were used only to identify the onset of EHIs. Patients were monitored for as long as they had combined ECG and blood pressure monitoring. The hospital utilizes GE Healthcare’s Carescape B850 and B650 monitoring systems (GE Heath Care, Chicago IL). NIBP measured by these systems uses automated oscillometric methodology.

Since the data was not specifically collected for this analysis and given the large dataset, no formal apriori power analysis was performed for this study. We estimated sensitivity, specificity, and other related measures for all AHI-PI outputs across NIBP and IAP monitored patient groups. For the purposes of the classification analysis, low and moderate risk AHI-PI outputs were grouped and evaluated against high-risk outputs. To evaluate the performance of the moderate risk class individually, risk ratios of an EHI for both moderate and high-risk outputs compared to low-risk outputs were calculated. Confidence intervals are also provided for each of the performance measures based on 1000 bootstrap samples with patient level replacement. This window-level analysis compares annotation of presence or absence of EHIs within a 1-hour prediction time frame against the AHI-PI scores for each window.

The lead time analysis assesses the question of ‘how far in advance does AHI-PI repeatedly produce a high-risk indication prior to an event?’ Therefore, using the onset of the first hemodynamic instability episode for a patient and looking back in time toward the beginning of AHI-PI monitoring, the duration of consecutive AHI-PI red (high-risk) outputs immediately prior to the onset of the EHI is calculated as the lead-time for that EHI (Figure 1). As some patients can have more than one EHI during their hospital stay and to avoid complications of counting multiple episodes and associating those episodes with specific AHI-PI high risk indicators, only lead times associated with the onset of the first EHI during AHI-PI monitoring were considered in calculating lead-time statistics. This provides a conservative measure of AHI-PI’s ability to predict the onset of such episodes. Given the nature of this assessment, lead times were only computed on the subset of patients who experienced at least one episode of hemodynamic instability.

**Figure 1:**
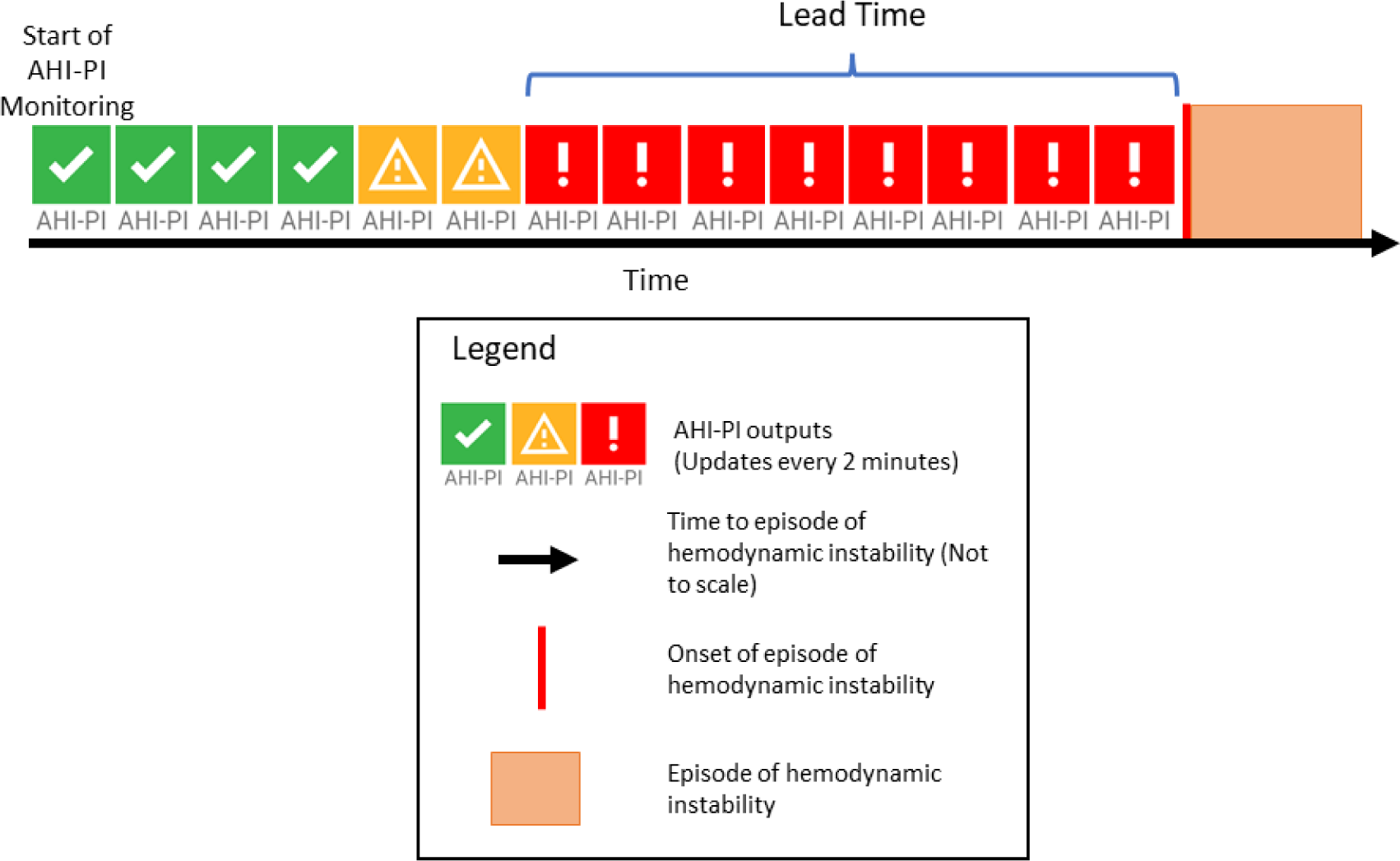
AHI-PI Lead time analysis approach. See text for details

Data analysis was performed using MATLAB 2021a (Natick, MA) to assess the test characteristics of AHI-PI including sensitivity, specificity, positive predictive value (PPV), negative predictive value (NPV), and the resulting receiver operator areas under the curve (AUC). Differences of AHI-PI outputs between the groups were analyzed using non-parametric (Wilcoxon Rank Sum Test) as well as parametric (Student’s t-test) methods.

## Results

Figure 2 demonstrates the breakdown of captured patients and data. Of 4,633 patients, 14.5% (672) had IAP monitoring. The other 85.5% (3,961) had NIBP monitoring with blood pressure verified by nurses and imputed into the EHR. Of all IAP monitored patients, 46.4% (312) had one or more EHI, while only 9.6% (380) of the NIBP monitored patients had one or more EHI.

**Figure 2:**
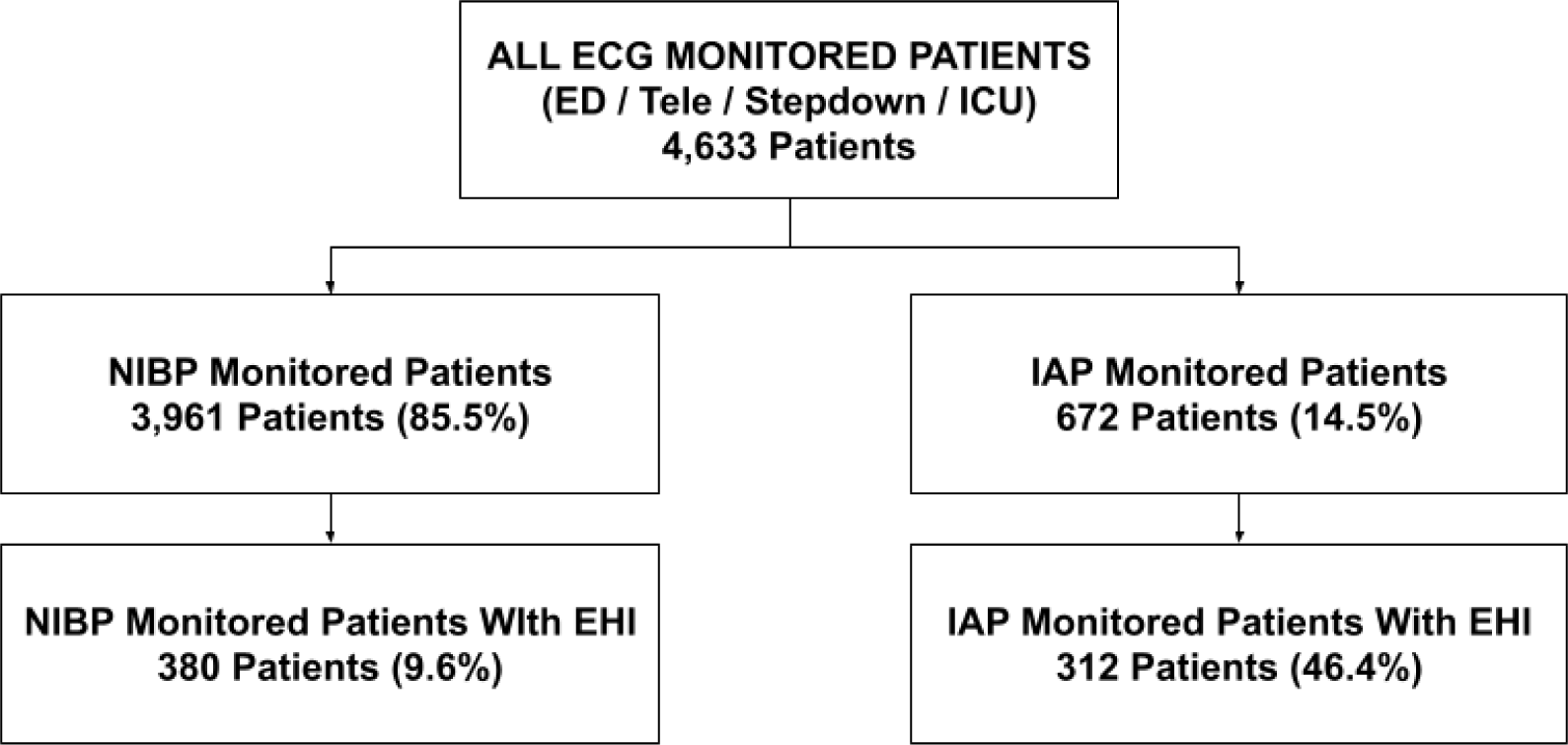
Consort diagram indicating study population and groups. NIBP: noninvasive blood pressure. IAP: intraarterial blood pressure.

Table 1 provides the demographic characteristics of the study population groups. The demographic and baseline characteristics were found to be similar between each of the groups. The average age in the patient groups ranged between 60.4 to 61.4 years, with the sex distribution slightly weighted towards males. The racial and ethnic distribution in the study population reflects the distribution of patients generally seen at the University.

**Table 1:**
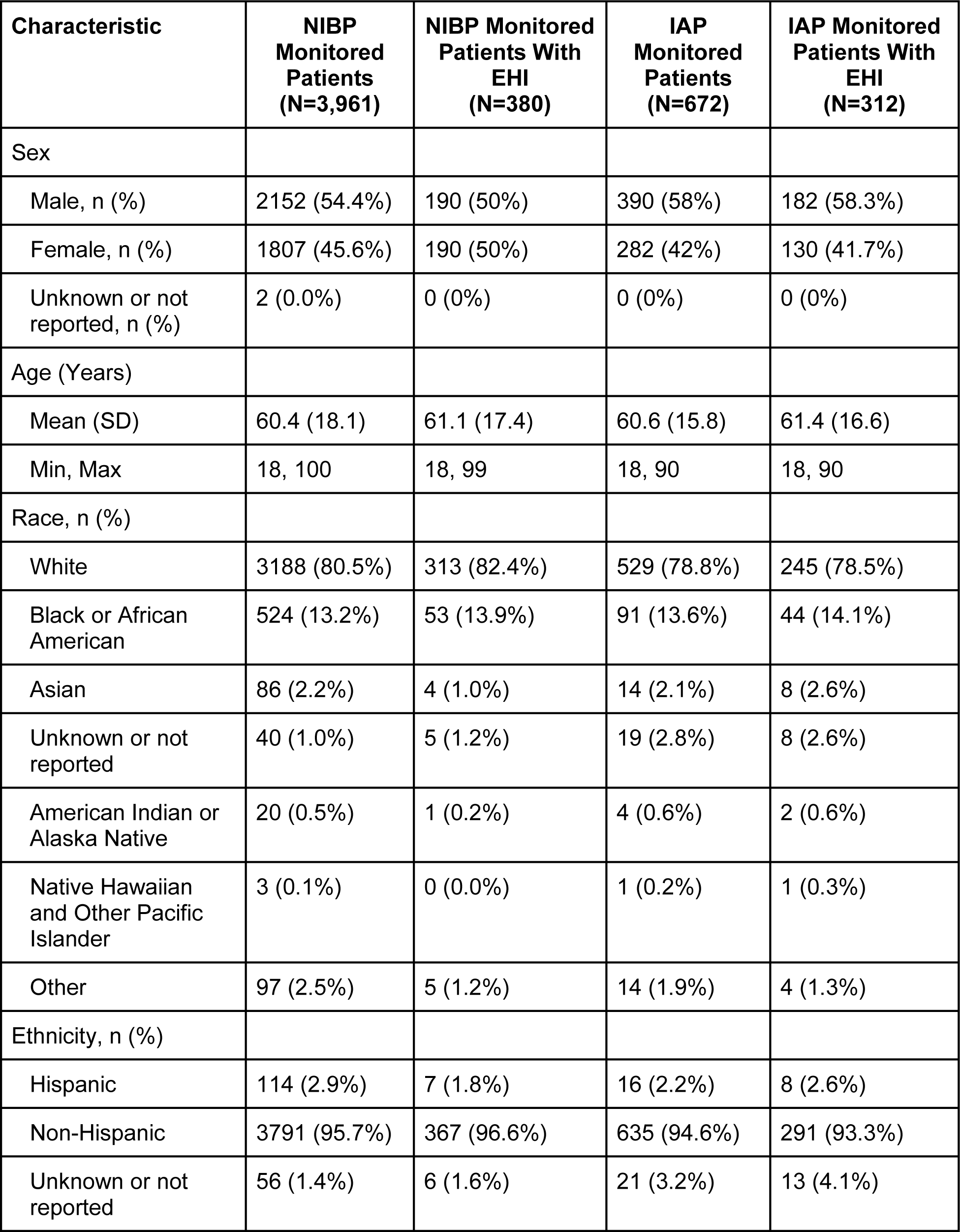
Demographic characteristics of the study population groups.

Table 2 provides the number and percentages of AHI-PI outputs based on low-risk, moderate-risk, and high-risk categories across both NIBP and IAP groups. Using all incidences of EHI across the three risk indicators, the risk of an EHI in the upcoming hour is 0.9% for IAP patients and 0.2% for NIBP given a low-risk AHI-PI output. Compared to this, patients demonstrating a moderate-risk output had a 6.2 (NIBP) and 6.7 (IAP) fold increased risk for an EHI in the next hour compared to a low-risk indicator. Finally, those patients having a high-risk output demonstrated a 35.7 (NIBP) and 38.9 (IAP) fold increased risk for an EHI compared to a low-risk indicator. This data formed the basis for combining the low- and moderate-risk categories to allow for performance measures.

**Table 2:** Number and percentage of AHI-PI Outputs based on risk categories.

| **AHI-PI Output Categories** | **Number (percentage) of Outputs** |
| --- | --- |
| **NIBP Low-risk** | 1,959,756 (69) |
| **NIBP Moderate-risk** | 270,285 (9.52) |
| **NIBP High-risk** | 610,088 (21.48) |
| **IAP Low-risk** | 914,659 (61.4) |
| **IAP Moderate-risk** | 139,937 (9.4) |
| **IAP High-risk** | 434,770 (29.2) |

AHI-PI’s sensitivity and specificity was 86.3% and 80.5% respectively for the NIBP monitored group and 89.7% and 78.3% respectively for the IAP monitored group, with an AUC of 0.87 for both groups (Table 3). PPV and NPV were 11.7% and 99.5% respectively in the NIBP group and 33.7 and 98.4% respectively in the IAP group. All data is presented in Table 3. Note that while the NIBP subset contained nearly 6x more patients than the IAP subset, there were only 2x the number of observations for classification-based measures. This is the result of more infrequent vital signs recordings in the NIBP population than IAP (once per hour vs 0.5 per second respectively).

**Table 3:** AHI-PI model performance measures in predicting presence or absence of Episodes of Hemodynamic Instability (EHI):

| **AHI-PI Window level analysis** | **NIBP Patients [95% CI]** | **IAP Patients [95% CI]** |
| --- | --- | --- |
| **Total Number of Patients** | 3,961 | 672 |
| **Number of AHI-PI Outputs - Low and High Risk** | 2,840,129 | 1,489,366 |
| **Prevalence** | 2.9% [2.5%, 3.4%] | 11.0% [9.7%, 12.3%] |
| **Sensitivity** | 86.3% [83.8%, 88.5%] | 89.7% [88.2%, 91.0%] |
| **Specificity** | 80.5% [78.8%, 82.2%] | 78.3% [75.7%, 80.6%] |
| **AUC** | 0.87 [0.85, 0.88] | 0.87 [0.86, 0.88] |
| **Positive Predictive Value** | 11.7% [9.5%, 14.0%] | 33.7% [30.1%, 37.3%] |
| **Negative Predictive Value** | 99.5% [99.4%, 99.6%] | 98.4% [98.0%, 98.7%] |
| **False Positive Rate** | 19.5% [17.5%, 21.6%] | 21.7% [19.0%, 24.9%] |
| **False Negative Rate** | 13.7% [11.1%, 16.8%] | 10.3% [8.8%, 12.1%] |

EHI predictions within a 1-hour prediction time frame against the AHI-PI scores for each window. Low and moderate risk groups are combined.

In the NIBP monitored patients with EHI, AHI-PI high-risk indications preceded EHI in 81.1% (308/380) of cases, with a median lead time of 1.1 hours (64 minutes) and average lead time of 2.9 hours (Table 4). Similarly in the subgroup of patients undergoing IAP monitoring (312), AHI-PI high-risk indications preceded the first EHI (10 continuous minutes of tachycardia and hypotension) in 83.3% (260/312) of cases, with a median lead time of 1.1 hours (66 minutes) and average lead time of 3.7 hours.

**Table 4:**
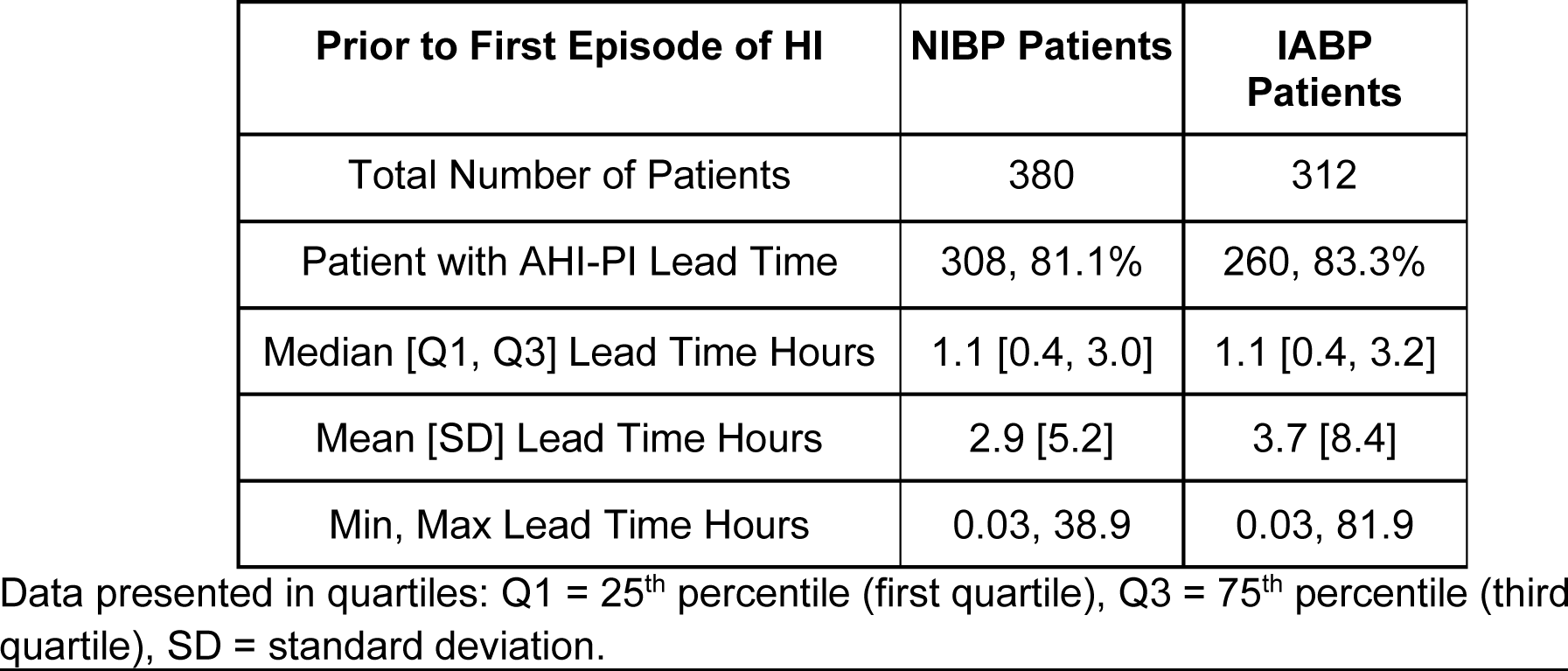
AHI-PI lead time to first episode of hemodynamic instability EHR Intended Use Population.

Table 5 provides the percent of AHI-PI high-risk outputs indicated in each of the hour-long periods prior to the first EHI across ECG monitored patients with an EHI. AHI-PI demonstrated a strong indication of future risk, with 89.7% or higher of the AHI-PI outputs in each hour-long period indicating high-risk, going back to two hours from the onset of the first EHI. Additionally, the distribution of percentage of AHI-PI between patients with and without EHI within the NIBP monitored patient group (3,961) is shown in Figure 3. The difference of AHI-PI high-risk outputs between the two groups was found to be statistically significant by both non-parametric and parametric testing with a p-value < 0.0001. The group of patients experiencing an EHI had close to 40% of their outputs indicating AHI-PI high-risk, compared to a median of 0% in the group of patients who did not experience an EHI. This indicates that AHI-PI predominantly indicates high-risk outputs in patients who experienced an EHI.

**Figure 3:**
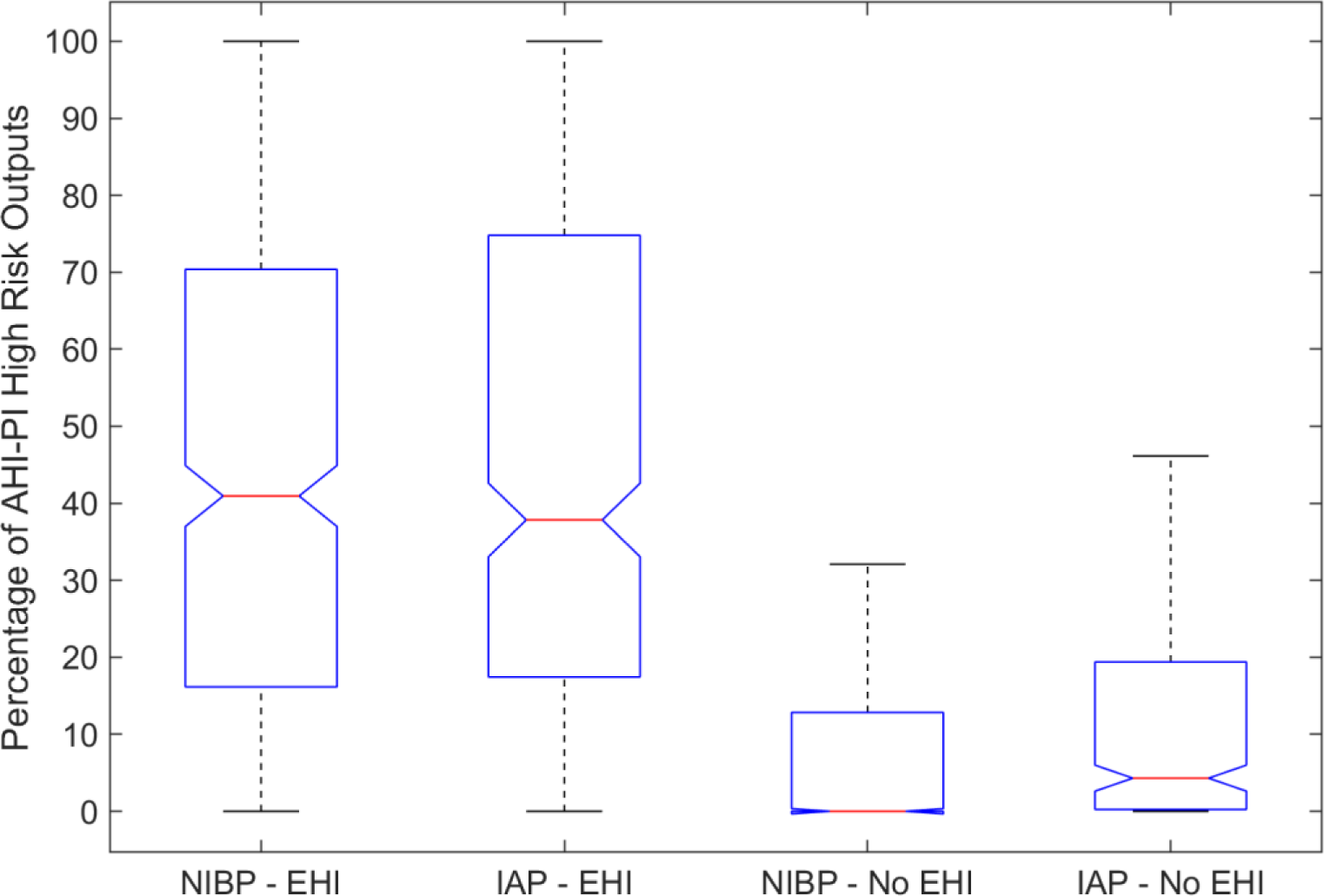
Distribution of AHI-PI High Risk indication – four groups of patients (patients with and without EHI). Patients monitored with IAP, NIBP. On each box, the central mark (red) indicates the median, and the bottom and top edges of the box indicate the 25th and 75th percentiles, respectively. The whiskers extend to the most extreme data points which are no longer than 50% greater than the interquartile range.

**Table 5.** Prevalence of AHI-PI High Risk Indications in the hours prior to the onset of the first EHI across all patients (NIBP and IAP monitored).

| Statistic | Monitoring Modality | 0-1 hours | 1-2 hours | 2-3 hours | 3-4 hours | 4-5 hours |
| --- | --- | --- | --- | --- | --- | --- |
| N patients | NIBP | 380 | 315 | 264 | 234 | 216 |
|  | IAP | 312 | 255 | 220 | 193 | 171 |
| Median [25th, 75th] | NIBP | 100%<br>[46.7%, 100%] | 89.7%<br>[0%, 100%] | 69.1%<br>[0%, 100%] | 48.8%<br>[0%, 100%] | 45.4%<br>[0%, 100%] |
|  | IAP | 100%<br>[43.3%, 100%] | 90.0%<br>[0%, 100%] | 61.7%<br>[0%, 100%] | 26.7%<br>[0%, 100%] | 0%<br>[0%, 100%] |
| Mean [Std Dev] | NIBP | 74.2%<br>[38.9%] | 58.8%<br>[45.1%] | 54.3%<br>[45.8%] | 48.8%<br>[46.2%] | 45.4%<br>[45.7%] |
|  | IAP | 72.3%<br>[39.2%] | 58.5%<br>[45.2%] | 53.1%<br>[46.1%] | 45.9%<br>[46.5%] | 40.1%<br>[46.1%] |

## Discussion

In this retrospective study we examined the utility of applying the AHI-PI analytic to detect an EHI using a cohort of patients who have ECG continuously collected along with either blood pressure continuously by IAP or intermittently collected and recorded by NIBP. AHI-PI demonstrated the ability to predict an EHI with high sensitivity and specificity with lead times that can be considered clinically relevant.

Predicting acute hemodynamic deterioration or instability in a time window to allow for intervention both in the intensive care unit and general ward setting continues to present a significant challenge. While various EWSs and scores exists, including but not limited to the Modified Early Warning System (MEWS), the National Early Warning System (NEWS), the Simplified Acute Physiology Scores (SAPS), and commercial products such as the EPIC’s deterioration index (EDI), none rely on continuous real-time analysis of one or more continuously collected physiologic variables.^4–6^ Instead, each are computed intermittently based on available structured data as it is imputed in the EMR. Consequently, they are contingent on the frequency of vital signs or laboratory data recorded in the EMR. As such, patients may develop physiologic changes and deterioration that occur in the interim or have causes for their hemodynamic deterioration that are not detected by the data collected from a particular EWS. Even in the ICU or operative setting, where dense high resolution physiologic signals are available, their continuous processing and analysis to predict acute adverse events such as hemodynamic instability is limited.

Other approaches and technologies have used hemodynamic waveforms to predict hemodynamic instability and hypotension. These include the Compensatory Reserve Index (CRI: Impact Vitals, Boston, MA), which uses changes in the photoplethysmograph (PPG) waveform associated with cardiac output and vasomotor changes to detect decompensation.^23, 24^ The Acumen Hypotension Predictive Index system (HPI, Edwards Lifesciences, Irvine CA) uses features in the arterial waveform (collected from IAP monitoring) to predict the occurrence of hypotension during surgery.^25, 26^ However, these approaches require separate hardware. AHI-PI is an SaMD that identifies HRV features associated with hemodynamic instability and is intended to be scaled to any monitor used for ECG monitoring. Its only requirement is a sampling rate of at least 120 Hz.

For AHI-PI as well as for any EWS, deterioration index, and clinical decision-making tool, there are important implications for errors. False negative outputs will mean care providers will see an AHI-PI output indicating stability when the patient is heading towards instability. If hemodynamic instability progresses unrecognized, there is the risk of failure to intervene resulting in an actual episode of instability. The high sensitivity reported in this study helps to minimize the potential to miss impending EHIs. Conversely, a false positive means the clinician will observe an AHI-PI unstable output when the patient is in fact stable and not progressing towards instability. The consequence of this can be the added resources required for increased vigilance and must be weighed in comparison to a false negative or when vital sign monitoring and reporting is less frequent. Since the AHI-PI high-risk outputs (true positives and false positives) are mainly concentrated in the group who do experience one or more EHI (Figure 3), AHI-PI may be viewed as having low alarm fatigue implications in the group without EHI (median 0% AHI-PI high-risk Indication). Since AHI-PI is intended as adjunctive data, which may be considered by care providers in determining types of care to be initiated, continued, or discontinued, the AHI-PI is envisioned to assist in clinical decision making.

PPVs between the two groups show a difference, reflecting the influence of the differences in prevalence between the two groups.^27^ As might be expected, the prevalence of hemodynamic instability is higher in the IAP group (11.0%) as compared to the NIBP monitoring group (2.9%). AHI was previously reported as being able to detect current hemodynamic status as opposed to predicting it^16^. The predictive aspect of AHI-PI provides a forward-looking risk assessment that supports clinical decision making. AHI-PI uses the same single lead of ECG (II) as input and combines AHI outputs that detect signs of hemodynamic instability into a single indicator that estimates the likelihood of a future adverse cardiovascular event or condition.

We believe there is a need for technology that allows for a more robust analysis of an easily obtained physiologic signal such as ECG that can be continuously analyzed to provide insight into a patient’s current and future physiologic state. We chose the ECG signal because of its ability to leverage HRV as a “vital sign” for the autonomic nervous system and its association with states of deterioration, and because of its ubiquity as a monitored signal. An additional advantage is it is much less conditioned by monitor manufacturers compared to other real-time signals and can now be monitored using a variety of adhesive patches designed for mobile monitoring. These patches allow monitoring of the ECG signal at resolutions and frequencies that provide HRV that are equivalent to traditional hospital-based monitors. The potential to use and scale such monitoring approaches may allow for more accurate triage and disposition of patients to or from higher or lower levels of care, including to general wards and even home, resulting in improved resource allocation. In addition, AHI-PI can be envisioned to guide treatments that move a patient from high-risk to low-risk status.

There are several limitations to this study. This includes the retrospective nature of the study and that the data utilized in this study comes from a single U.S. academic health center. We also used previous but well-defined definitions of stability and instability based on vital signs.^4, 20– 22, 28^ While instability characterized by only hypotension or only tachycardia can occur, we sought to use a more robust definition that combines these vital signs, which are more likely to indicate both issues with circulatory perfusion (hypotension) and the burden on the autonomic nervous systems through sympathetic activation (tachycardia), which may lead to compensatory failure and shock if left unstreated.^29^ Changes in these criteria would result in the need for more analysis of the performance of AHI-PI.

It should be recognized that patients who were on a trajectory for an EHI as identified by AHI-PI may have undergone various treatments that prevented the EHI before it occurred. Patients with cardiac transplant, ventricular assist devices, sustained atrial or ventricular arrythmias, or pacemaker dependence were not excluded. These conditions could adversely impact HRV as a measure of the autonomic nervous system. Future studies that control for these issues may result in improved performances of AHI-PI. We also acknowledge the reported inaccuracies of noninvasive oscillometric blood pressure monitoring when compared to intra-arterial blood pressure monitoring in acute care patients.^30–33^ AHI-PI in this setting may have been potentially more accurate given we paired it as a continuous measure with a significantly sparser oscillometric based NIBP measurement.

Lastly, we did not explore the reason or potential reason for patients’ EHI, what actions were undertaken to treat it, or complications from the EHI. This and the other limitations above are topics of ongoing studies.

## Conclusion

Accurate prediction of an EHI may allow for improved resource allocation, shorter time to intervene, changes in disposition, and better patient outcomes. However, such predictions are challenging when using intermittent vital signs. This study supports the potential use of a novel ECG monitoring strategy and analytic that leverages signal processing and machine learning to extract ECG features associated with impending hemodynamic instability. The noninvasive nature of the technology may offer advantages in continuous surveillance and real-time clinical decision making, facilitating interventions to prevent EHIs.

## Data Availability

The datasets generated and analyzed in this study are not publicly available due to restrictions in place by both the institutional IRB under which the study was performed as well as a data use agreement between the University and Fiftheye Inc. Limited data sharing is available from the corresponding author on reasonable request.

## Abbreviations

AHI-PI: Analytic for Hemodynamic Instability-Predictive Indicator
AUC: area under the curve
CRI: Compensatory Reserve Index
eCART: electronic Cardiac Arrest Triage
ECG: electrocardiogram
EDI: EPIC’s Deterioration Index
EHI: episode of hemodynamic instability
EMR: electronic medical record
EWS: early warning systems
HPI: Hypotension Predictive Index
HRV: heart rate variability
IAP: intra-arterial pressure
ICU: intensive care unit
MEWS: Modified Early Warning Score
NEWS: National Early Warning Score
NIBP: noninvasive blood pressure
NPV: negative predictive value
PPG: photoplethysmograph
PPV: positive predictive value
SaMD: software as a medical device
SAPS: Simplified Acute Physiology Scores

## Declarations

### Ethics approval and consent to participate

This study was approved by the University of Michigan’s Institutional Review Board. Due to the retrospective nature of the study and the use of de-identified data, the Institutional Review Board waived the requirement for patient informed consent.

### Informed Consent

The University of Michigan’s Institutional Review Board and its Human Research Protection Program provided a waiver of informed consent for this study due to its retrospective design and use of de-identified data.

### Guidelines

All methods described in this study were carried out in accordance with relevant guidelines and regulation in accordance with the Declaration of Helsinki.

### Consent for publication

Not applicable

### Competing and Conflicts of Interest

Ashwin Belle and Kevin R. Ward are inventors of the AHI technology with patents assigned to the University of Michigan and Fiftheye Inc. Bryce Benson, Ashwin Belle, Sooin Lee and Kevin R. Ward have equity in Fiftheye Inc. Benjamin S. Bassin, Richard Medlin, and Michael W. Sjoding have no competing interests.

### Funding

Not applicable

### Authors’ contributions

Bryce Benson, Ashwin Belle, Benjamin S. Bassin, and Kevin R. Ward conceived and designed the study. Bryce Benson, Ashwin Belle, Sooin Lee, Richard Medlin, Michael W. Sjoding, and Kevin R. Ward analyzed data. All authors participated in the manuscript development and review.

## Acknowledgements

The authors would like to thank Mr. Santinio Jones for his assistance in manuscript preparation and submission.

## Authors’ information (optional)

Not applicable

## Footnotes

Not applicable.

## Notes

### Author Declarations

This study was approved by the University of Michigans Institutional Review Board. Due to the retrospective nature of the study and the use of de-identified data the Institutional Review Board waived the requirement for patient informed consent.

